# Transdiagnostic phenotyping of psychopathology in a help-seeking population (PhenoNetz): A study protocol for an experience sampling study

**DOI:** 10.1101/2021.05.10.21256804

**Authors:** Marlene Rosen, Linda T. Betz, Christian Montag, Christopher Kannen, Joseph Kambeitz

**Affiliations:** Department of Psychiatry and Psychotherapy, Faculty of Medicine and University Hospital of Cologne, University of Cologne, Cologne, Germany; Institute of Psychology and Education, Ulm University, Ulm, Germany

**Keywords:** help-seeking population, phenotyping, ecological momentary assessment, symptom networks, prevention, early intervention, transdiagnostic psychiatry

## Abstract

Prevention in psychiatry provides a promising way to address the burden by mental illness. However, established approaches focus on specific diagnoses and do not address the heterogeneity and manifold potential outcomes of help-seeking populations that present at early recognition services. Conceptualizing psychopathology manifested in help-seeking populations from a network perspective of interacting symptoms allows transdiagnostic investigations beyond binary disease categories. Furthermore, modern technologies such as smartphones facilitate the application of Experience Sampling Methods (ESM). A combination of ESM with network analyses provides valid insights beyond established assessment instruments. We will examine n = 75 individuals (age 18-40 years) of the help-seeking population of the Cologne early recognition centre (FETZ). For a maximally naturalistic sample, only minimal exclusion criteria will be applied. We will collect data for 14 days utilizing a mobile application to assess ten transdiagnostic symptoms, i.e., depressive, anxious and psychotic symptoms as well as distress level. With these data, we will generate average group-level symptom networks and personalized symptom networks using a two-step multilevel vector autoregressive model. Additionally, we will explore associations between symptom networks and sociodemographic, risk and resilience factors, as well as psychosocial functioning. Our study will provide insights about feasibility and utility of ESM in a help-seeking population. Providing a first explorative phenotyping of the transdiagnostic help-seeking population, this study will contribute to innovation of early recognition in psychiatry. Results will help to pave the way for prevention and targeted early intervention in a broader patient group and thus, enable greater intended effects in alleviating the burden of psychiatric disorders.

## Introduction

Prevention and early intervention in psychiatry provide promising ways to address the immense burden of mental illness [1–3]. The currently established prevention approach implemented in early recognition services focuses on risk syndromes developed for predicting specific diagnoses (e.g., psychosis [4,5]). However, the majority of help-seeking patients who present at early recognition services are not covered by these specific risk syndromes, as they do not fulfill respective criteria which indicate increased risk qualifying for targeted intervention [4,6]. Thus, in a sizable proportion of this population, early recognition centers for mental disorders currently miss out on a critical potential for preventive efforts. In fact, help-seeking populations present with a mixture of various symptoms [7] such as depressive, anxious and psychotic symptoms. Depressive and anxiety symptoms have proven to be among the main reasons why individuals seek help [8,9], whereas psychotic symptoms are of interest as they are most burdensome for affected individuals as well as for the health care system, despite their rather low prevalence [10].

These symptoms are shared across different diagnoses [11–13] and appear in help-seeking patients with sub-threshold, risk-syndromes and full-threshold disorders [14]. Similarly, accumulating evidence demonstrates that distress is a mediating and triggering factor for psychopathology at large [15–19]. Taken together, help-seeking populations are much more heterogeneous than previously assumed, and may develop manifold potential outcomes [20] or show other unfavorable outcomes such as persisting deficits in psychosocial functioning [21].

Thus, there is a growing call for a broader, transdiagnostic approach for prevention in psychiatry [22–25]. While there is important data on the psychopathology of patients presenting to early recognition centers (e.g. [6,8,26,27]), their interpretation is limited by the typically purely cross-sectional, retrospective and diagnosis-specific character of the assessments. However, symptoms fluctuate over time [28–30], and important insights are missed when neglecting this dynamic component of psychopathology in a help-seeking population. Moreover, as outlined above, conventional assessments are often considered in isolation rather than in concert, neglecting the transdiagnostic, intertwined nature of psychopathology in help-seeking populations. Collectively, these observations underline the necessity to turn to novel methods in order to enrich traditional self- and observer-based reports to understand the psychopathology in help-seeking populations.

One novel method consists in integrating two distinct innovative ideas that have emerged in the field of psychopathology in recent years [31]. The first consists in intensive longitudinal measurements of symptoms and other relevant variables via the experience sampling method (ESM), which has become increasingly feasible and accepted in recent years especially with the advance of mobile technology such as smartphones (see [32] and [33]). ESM provides valid insights into psychopathology as it occurs in daily life by assessing targeted phenomena repeatedly during the course of the day within a specific time period. ESM increases ecological validity compared with retrospective reporting, reduces biases resulting from false memory or aggregation processes of experience over a longer time period, and allows collection of data at the within-person level [34].

The second idea is the network approach mainly put forward by Borsboom and colleagues ([35–37]; for a recent overview, see [38]), in which psychopathology is conceived as a dynamic system of connected, interacting and maintaining symptoms and other clinically relevant variables [35,39]. In line with a clinician’s perspective, symptoms are assumed to co-occur because of functional relations between them rather than due the common dependence on an underlying disorder entity [36,38,39]. With its inherently transdiagnostic outlook [37,40], the network approach is well suited for conceptualizing the psychopathology of a help-seeking population, where patterns and strength of symptom expression is typically highly heterogeneous.

The integration of network analyses with ESM data enables rich insights beyond those obtained by established assessment instruments. Specifically, the intensive time-series data the result from ESM can be used to model symptom networks that offer a promising gateway into understanding the dynamics of psychopathology on group and individual level [41]. On group-level, dynamic symptom networks allow to exploratively map out the potential average causal relations among individual symptoms in the same measurement window and across measurement windows. Personalized symptom networks are of special interest, as they allow the conceptualization of psychopathology as a set of person-specific dynamic processes [39,42]. By revealing the symptoms and processes most relevant to each individual, these approaches hold potential to personalize interventions [39,43].

Due to these properties, many interesting studies have been published proving the potential of longitudinal symptom network models in advancing the psychopathological understanding of specific psychiatric conditions [44,45]. However, insights into the dynamical structure of psychopathology of a heterogeneous help-seeking population of a psychiatric early recognition center, i.e., interactions of a broad, transdiagnostic set of symptoms, as well as associations with risk and resilience factors and psychosocial functioning, is still lacking so far.

Thus, with the proposed study, we aim to provide a first explorative transdiagnostic phenotyping through the combination of ESM with network analyses. This will be the first study to apply ESM in a help-seeking population.

Findings from this innovative approach integrated with those derived from established assessments represents a promising way to address a larger proportion of the help-seeking population than current diagnosis-specific strategies aiming at preventing the burden of psychiatric disorders. Moreover, results will have their core value in generating hypotheses regarding central dynamic psychopathological processes. These provide a basis for follow-up work dedicated to inform preventive interventions, testing experimentally whether interventions on particular symptoms or processes lead to changes consistent with estimated network model [46].

## Methods

### Aim

This study aims at an explorative transdiagnostic phenotyping of a help-seeking population of an early recognition center for mental disorders using innovative, intensive longitudinal data collection via a smartphone app. A better understanding of relevant psychopathology in this burdened population is of great relevance, given the lack of adequate interventions [47]. Combining ESM with network analyses allows for unique insights into yet under-researched early transdiagnostic psychopathological processes in a help-seeking population of an early recognition center of mental disorders, as well as their explorative association with risk, resilience, and psychosocial functioning.

### Setting and participants

100 participants will be recruited from the help-seeking population presenting at the early recognition center of mental disorders at the University Hospital of Cologne (FETZ) (fetz.uk-koeln.de), with an expected drop-out rate of 25%, leading to a total of 75 participants in the final sample. The FETZ offers specialist diagnostics for the early recognition of mental disorders, with a focus on severe mental illness, in particular psychotic disorders, and is a first contact point for people aged 18 to 40 years that have noticed changes in their experience and behaviour. Most patients find out about the FETZ through internet research or are referred to by healthcare practitioners.

For a naturalistic characterization of the help-seeking population presenting at the FETZ using ESM, we will not impose specific inclusion criteria for participation in the PhenoNetz-study. Likewise, to ensure validity of the obtained data, only a minor part of help-seeking participants will be excluded based on the following criteria:

- acute suicidal thoughts
- IQ ≤ 70
- age > 40 years
- known previous illness of the central nervous system, as well as untreated unstable somatic illnesses with known effects on the central nervous system (e.g. untreated hypothyreosis)
- insufficient knowledge of the German language

### Procedure and materials

All patients presenting at the FETZ not fulfilling any of the listed exclusion criteria will be addressed either directly in the FETZ or via telephone or e-mail (given permission to contact was obtained by the clinical personnel at the FETZ) and informed about background, goal, design, risks and benefits, as well as data security aspects of the study. Additional open questions will be answered directly by one of the primary investigators (MR, LB). In case of willingness to participate, written informed consent will be provided by all participants prior to their participation in the study. All participants will be compensated with 40€ for their participation. Participants can withdraw from the study at any time without negative consequences. The study was approved by the Institutional Review Board of the University of Cologne, Faculty of Medicine (reference number 20-1092).

Figure 1 illustrates the study design. During baseline assessment, data on socio-demographics, medication, substance use, psychopathology including psychosocial functioning as well as risk- and resilience factors will be assessed through both observer- and self-ratings (table 2). All data will be collected via Research electronic data capture (RedCap [48]). In the baseline assessment, the mobile application used for ESM data collection in the study, the *insightsApp* [49], will be installed on the personal smartphones of the participants. As the *insightsApp* only runs on Android, participants with personal smartphones using other operating systems (e.g., iOS) will be equipped with a study smartphone for the study period. Participants will be encouraged to complete as many surveys as possible without compromising their personal safety (e.g., while driving).

**Figure 1.**
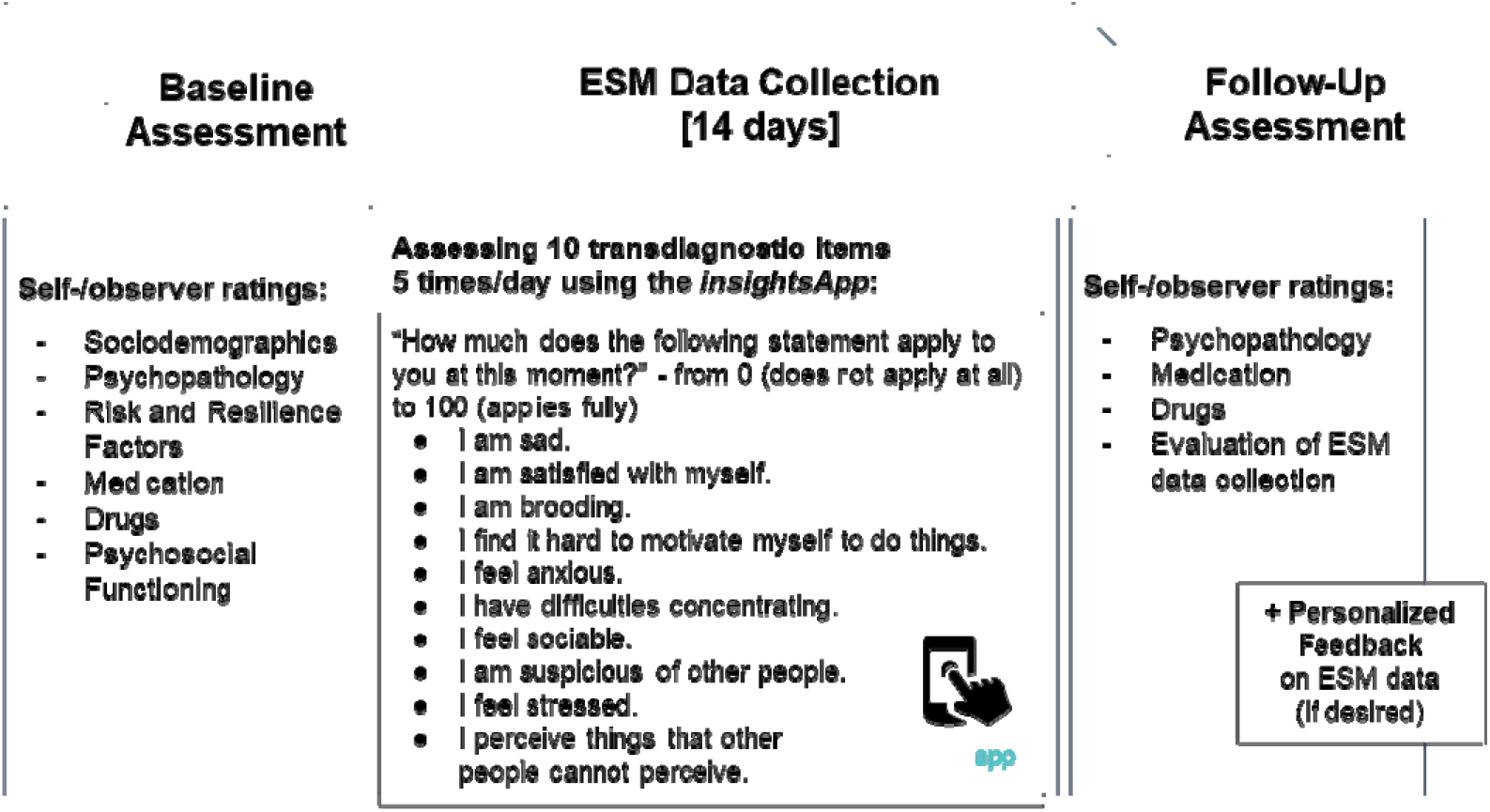
Study design of the PhenoNetz-study. Participants included will undergo baseline assessment with self- and observer ratings, followed by a 14-day-ESM data collection period. In the subsequent follow-up assessment, selected self- and observer ratings will be collected again. If desired, the participants will receive personalized feedback on their ESM data after the two weeks of ESM data collection, such that the feedback does not interfere with ESM data collection.

**Table 2.**
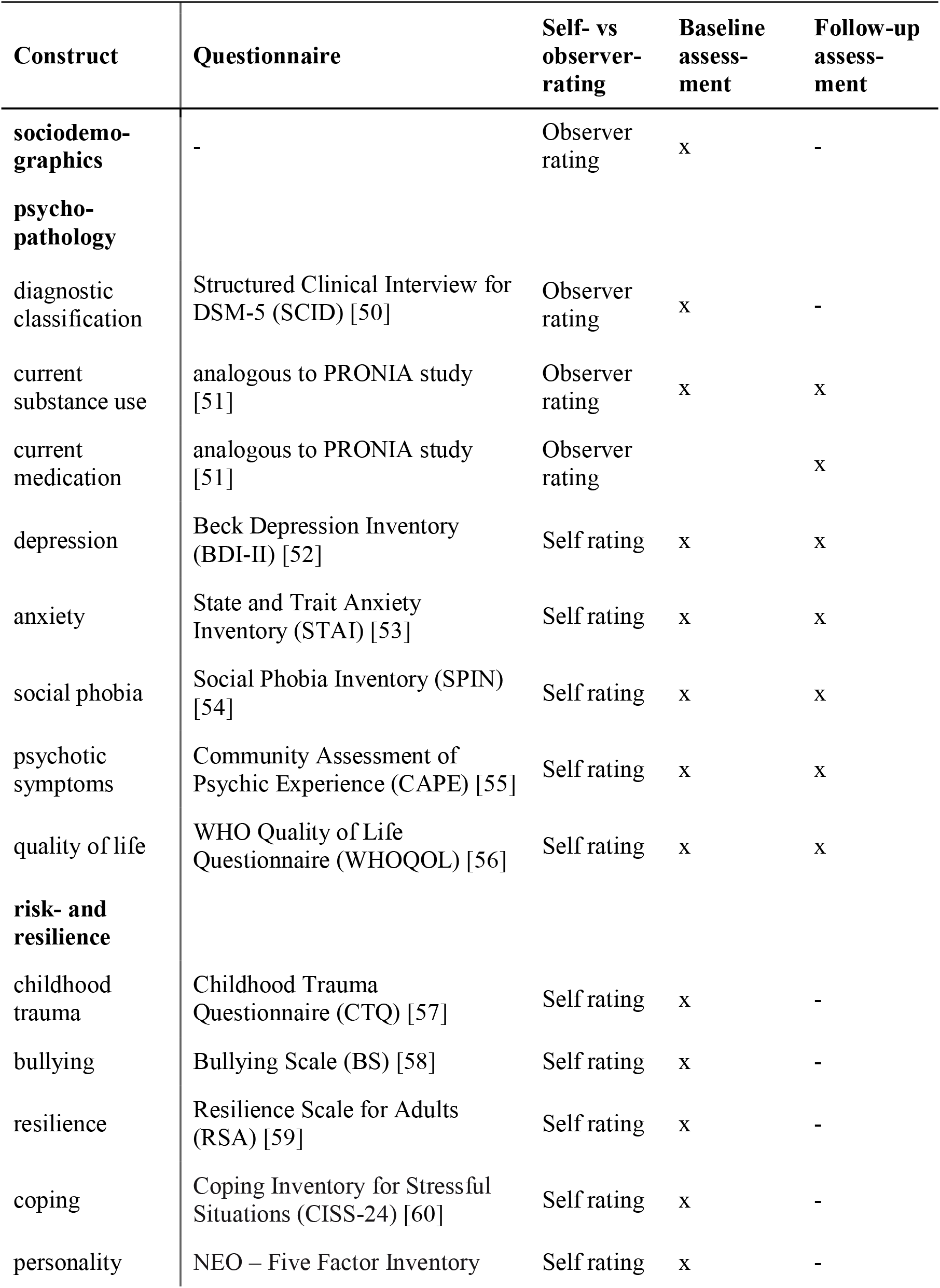
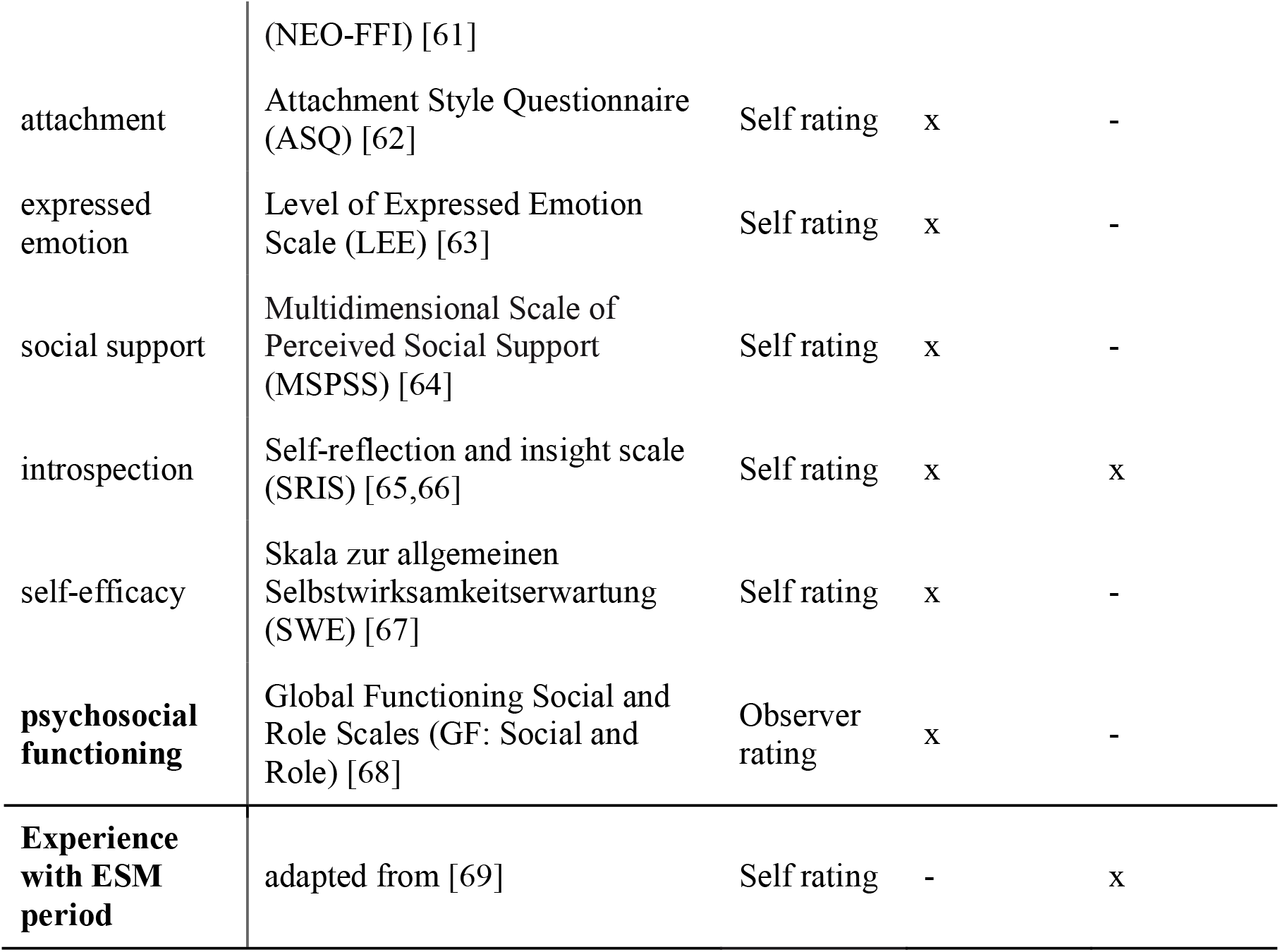
Constructs with scales assessed at the baseline and follow-up assessments (before and after the ESM period, respectively) of the PhenoNetz-study.

Using ESM, potentially relevant transdiagnostic (subthreshold) symptoms such as sadness, anxiety, psychotic experiences and stress will be recorded (table 3). Items are based on previous studies and questionnaires, given the lack of standardized ESM assessment in clinical populations. In-app reminders will be sent out five times a day at fixed time points: 9:30h, 12:30h, 15:30h, 18:30h, 21:30h for a duration of 14 days. In each survey, participants will be asked how much they endorse a certain feeling or behavior at the time of filling out the survey: “How much does the following statement apply to you at this moment?”. Responses will be given on a visual analogue scale (in percent) from “0 = does not apply at all” to “100 = “applies fully”, with a slider that can be moved in 1-unit increments. Participants will be asked to fill in the items as soon as possible after receiving the in-app reminder, but no later than 60 minutes afterwards. Filling in the items takes about 1-1.5 minutes in total. Similar ESM protocols were deemed acceptable for clinical populations in prior studies [34,69,70]. The *insightsApp* will be used only for the regular active collection of transdiagnostic symptoms by means of the described self-report questions. No personal information (such as name, phone number, etc.) or passive data are accessed, stored or transferred by the *insightsApp*. To maximize the number of completed surveys for every participant, participants will be contacted at least once during the assessment period to assess instruction adherence, identify any concerns associated with the method and help participants with any problems in completing the ESM questionnaire.

**Table 3.** ESM items assessed in the PhenoNetz-study (English translation).

|  |  |
| --- | --- |
| 1. | I am sad. |
| 2. | I perceive things that other people cannot perceive. |
| 3. | I have difficulties concentrating. |
| 4. | I feel sociable. |
| 5. | I feel stressed. |
| 6. | I am satisfied with myself. |
| 7. | I feel anxious. |
| 8. | I find it hard to motivate myself to do things. |
| 9. | I am suspicious of other people. |
| 10. | I am brooding. |

In the follow-up assessment conducted after the 14 days of ESM data collection, data on psychopathology, medication and substance use will be assessed again, referring to the 14 days during which ESM data were collected (table 2). In addition, experiences and strain associated with the ESM data collection will be assessed via a questionnaire translated and adjusted from a previous study conducted in clinical participants [69]. If desired, participants will be provided with a personalized feedback report on their ESM data.

### Data security

Using a smartphone app installed on the personal smartphone of participants for data assessment requires particular attention to data security (for a broader discussion on ethical concerns regarding digital phenotyping procedures in the psychological and psychiatric sciences see [71,72]). Therefore, subjects must provide additional consent to collect data within the app and grant the necessary permissions to the app on the smartphones (such as being notified by the app about available surveys). The ESM data collected by the *insightsApp* is pseudonymized (16-digit alphanumeric codes) and sent directly to a server hosted and maintained by a professional web hosting service after each survey. Answers to the surveys are only stored temporarily locally on the smartphones and deleted once they are transmitted to the server. To secure the data transfer from the smartphone to the server, both the connection between the *insightsApp* and the backend software on the server is encrypted by the use of a SSL certificate.

### Safety

Given that this study is observational, there are no direct risks associated with participation. Previous studies have demonstrated good acceptance of ESM protocols similar to the one implemented in this project. Even if participants become more aware of their symptoms due to high-frequency data collection, this does not have a negative effect in terms of worsening symptoms [34,69,73]. Participants can terminate the ESM data collection at any time without giving reasons. Participants who are acutely suicidal or a danger to others will immediately be presented to the service physician for further assessment. Should this become apparent in a telephone call, participants will be reported to the responsible social psychiatric service.

### Data analytic plan

All statistical analyses will be conducted in the *R* language for statistical computing [74]. Descriptive analysis of the sample will include mean, standard deviation, median and interquartile range as appropriate. Participants included in the analysis will be compared to those that dropped out of the study or were excluded due to too little available measurements (< 20 measurements [41,44]) via appropriate classes of permutation tests [75]. Changes in measures that were assessed twice, pre- and post-ESM (see table 2), will be compared via linear mixed modeling. Prior to the analyses of ESM data, we will detrend the ESM data by fitting fixed-effects linear regression models to each ESM item, regressing out a linear trend on time (i.e., general increases/decreases in items over time), and mean-center ESM items per person. We will then generate group-level and personalized networks via a two-step multilevel vector autoregressive (VAR) modeling approach as described in detail below. These analyses allow us to examine symptom dynamics within multiple individuals (*n* > 1; fixed effects) and for individual participants (*n* = 1; random effects). Originally, we planned to estimate and analyze ‘truly’ personalized networks solely based on data from individual participants (such as those that could be derived via a graphical vector autoregressive model [31]). However, results from a simulation study [76], published as a preprint one month after our study commenced, suggest that our sampling scheme potentially lacks the power to detect a non-negligible proportion of true edges in truly personalized networks, which is why we decided to refrain from this analytic approach.

### Two-step multilevel vector autoregressive model

We will use a two-step multilevel vector autoregressive (VAR) model, as implemented in the mlVAR *R* package. In the multilevel VAR, the average dynamical relationships on group-level are modeled as fixed effects, whereas regression coefficients are allowed to vary between patients as random effects.

First, we will estimate three group-level network structures including the 10 assessed symptoms, reflecting the average process of all participants (fixed effects): between-subject (undirected partial correlation network between the means of participant’s scores, capturing, in general, whether participants high on a given node are also high on other nodes during the two-week course of the study), contemporaneous (an undirected partial correlation network showing how symptoms relate to each other in the same window of measurement, controlling for temporal relationships), and temporal (a directed network displaying symptoms predicting each other across an approximately 3-h lag, while controlling for all other experiences in the model at the prior measurement). Centrality will be assessed using strength centrality (indicating the summed absolute edge strengths connected to a specific node) in the contemporaneous network, and in-strength (indicating the summed absolute strengths of all incoming edges) and out-strength (indicating the summed absolute strengths of all outgoing edges) in the temporal network, using the *R* package ‘qgraph’ [77].

Second, we will generate two types of personalized networks for each participant based on estimated random effects of the multilevel VAR: a contemporaneous and a temporal network. These personalized networks are not truly idiographic, in the sense that they borrow information from other subjects [41,44]. However, in doing so, the multilevel VAR can perform well in estimating personalized networks even if the number of ESM observations is comparatively low for a particular participant. Given that multilevel VAR does not perform participant-specific model selection, all estimated personalized networks contain all edges [41].

We will use orthogonal estimation for contemporaneous and temporal effects. For the contemporaneous and group-level networks, we will use the conservative “AND-rule” approach in retaining and plotting significant edges. See [41,44] for a detailed description of methodological details.

### Specific analyses

We plan the following analyses:

1. We will compute group longitudinal (between-person, contemporaneous, temporal) networks (contemporaneous and temporal; see [31]) as described above.
2. We will identify symptom centrality and unique partial correlations among symptoms in the contemporaneous and temporal group-level networks. We hypothesize that on group-level, feeling stressed will be the most central symptom in the contemporaneous network and predict other experiences most in the temporal network, given that stress experience is frequently discussed as a transdiagnostic factor in psychopathological experiences [16–19]. For the temporal network, we have no a priori hypothesis with regard to the most central item.
3. We will evaluate the degree of association between risk factors (e.g. childhood trauma) and network connectivity, assessed by global strength of personalized networks (temporal and contemporaneous), in a linear modeling approach. Based on prior research and theoretical considerations [36,78,79], we hypothesize that risk factors will be associated with an increased network connectivity. Similarly, we hypothesize that poorer psychosocial functioning will be associated with increased network connectivity.
4. We will explore how the strength of specific symptom-symptom connections in individual contemporaneous and temporal networks relates to the degree of presence of specific risk/resilience factors.

### Sample size and required number of ESM observations

Formal power analyses are not yet worked out for group-level network models based on intensive longitudinal data. The performance of network estimation methods depends on the unknown true network structure—the network equivalent of a true effect size in power analysis [44]. Supplementary materials from Epskamp, Waldorp, et al. (2018) report simulation results for mlVAR, showing that mlVAR models are excellent in recovering the fixed effect structures with quite few data, starting at 50 participants. With our targeted sample size of 75, which represents a realistic recruitment goal in the population of interest, we will surpass this threshold, leading to an adequately powered analysis for the estimation of a mlVAR model. Due to the methodological novelty of symptom networks based on intensive time series data, there exist no guidelines on the number of ESM observations required [44]. More observations collected over a longer period of time improves stability and validity of the results; however, this has to be balanced against the feasibility of the integration of the study into the daily lives of the participants. With 70 targeted observations collected over 14 days, our study is similar to the study designs of previous ESM projects conducted in clinical populations [80–82]. Following previous studies using mlVAR modeling, participants with less than 20 ESM observations will be excluded from the network estimation [41,44].

### Status and timeline of the study

Study recruitment started on November 11, 2020, and is currently ongoing, with an anticipated date of recruitment completion by the end of November 2021.

## Discussion

This study aims at an explorative phenotyping of the heterogenous help-seeking population of a psychiatric early recognition center. Applying ESM for the first time in such a cohort, we will identify transdiagnostic symptom networks and explore their association with protective and risk factors, as well as psychosocial functioning.

In doing so, we provide a first attempt to validly depict dynamic transdiagnostic symptom associations in a help-seeking population which might provide valuable insights: Central items and processes might represent anchor points for interventions [83]. Furthermore, insights into potential etiological processes, identified by association with risk and resilience factors as well as psychosocial functioning, might inform prevention strategies [47]. Hypotheses based on findings of our explorative study might guide future research.

Moreover, ESM represents only one powerful element to gain insights into relevant variables collected in everyday life to improve prevention and targeted early intervention [84,85]. Studying the digital footprints left by the human-smartphone interaction (e.g. log in frequency, use of different apps, calling behavior), can provide additional important insights into psychopathological states in help-seeking individuals [86]. Exploring the potential of ESM as a self-monitoring intervention in help-seeking populations (similar to approaches in depressive disorder [29]) is another exciting avenue for future research.

In clinical science, intensive longitudinal assessments of symptoms in daily life are deservedly receiving more and more attention [39,43,44], and we believe that this will result in enhanced patient benefit. Our study intends to contribute a milestone towards innovation in understanding help-seeking populations in psychiatry, allowing to help a greater proportion of this heterogeneous and crucial target group [43]. Subsequent impacts on early states and the progress of mental disorders might reduce associated personal, familial, societal, clinical as well as economic burden more effectively.

## Data Availability

The submitted manuscript is a study protocol and has no attached data.

## Acknowledgements

The study is funded by the Köln Fortune program, Faculty of Medicine, University of Cologne (grant agreement n° 304/2020).

## Multimedia Appendix of supplementary files

See attached file.

